# Remote Research Practices Enhance Acute Stroke Clinical Trial Enrollment

**DOI:** 10.1101/2023.02.17.23286102

**Authors:** Christopher D Streib, Abbey Staugaitis, Megan Tessmer, Denise Gaffney, Shayan Khan, Joseph P. Broderick, Pooja Khatri, Navdeep S Sangha

## Abstract

**Background:** Many acute stroke clinical trials (ASCTs) are underpowered or terminated early due to poor recruitment. A pervasive challenge to ASCT recruitment is the physical separation of patients, their legally authorized representatives, research coordinators, and clinician investigators when trial-eligible patients are identified. Remote research practices (RRPs) can facilitate time-critical ASCT enrollment and follow-up assessments, however, the feasibility and effectiveness are unknown.

**Methods:** This case-control study retrospectively reviewed ASCT enrollment at two institutions. When conventional in-person clinical research was not possible, completion of study specific essential clinical trial events (ECTEs) were attempted via RRPs utilizing telemedicine evaluation or telephone communication. The primary outcome was successful execution of ECTEs by modality: in-person, telemedicine evaluation, or telephone communication. The secondary outcome was protocol violation rate by modality. We utilized Fisher’s Exact Test for primary and secondary outcomes and descriptive statistics to report RRP utilization.

**Results:** A total of 1603 individual ECTEs were attempted for 171 subjects. RRPs were utilized for 53.9% of ECTEs (19.3% telemedicine, 34.6% telephone communication). ECTEs were more likely to be completed successfully via telemedicine (100%) than in-person (98.2%) or telephone (92.3%), (p<0.01). Additionally, protocol deviations were less common with telemedicine (0.0%) than in-person (2.6%), or telephone (2.8%) (p=0.04). More than half (53.4%) of randomized ASCT enrollments were dependent upon RRPs. Randomization (94.7%) and outpatient assessments (84.6%) were more frequently completed via RRPs compared to eligibility screening (40.7%), informed consent (40.4%), supervision of study intervention (44.6%), and inpatient assessments (18.8%).

**Conclusion:** Remote research practices were effective and doubled randomized ASCT enrollments in comparison to conventional research models that are restricted to in-person interaction alone. Telemedicine was associated with the highest rate of successful ECTE execution and the lowest rate of protocol deviations. These findings may be confounded by indication and further definitive study is indicated.

## Introduction

High quality clinical trials are essential to advance stroke care, however, many stroke clinical trials are underpowered, inconclusive, or terminated early due to poor recruitment.^1^ Recruitment challenges in acute stroke trials include rapidly identifying eligible subjects, determining trial eligibility, assessing patient capacity, obtaining and documenting informed consent from the patient or their legally authorized representative (LAR), and administering time-sensitive study interventions.^1^ The inability to complete these essential clinical trial events (ECTEs) hinders study enrollment and compromises the evaluation of promising new therapies.^2^ Furthermore, suboptimal completion of ECTEs could result in a number of adverse outcomes for the subject including: delaying effective standard of care treatment due to concomitant research activities, directly causing harm through the administration of contraindicated study interventions (e.g. thrombolytics), or violating patient autonomy through inadequate informed consent that does not accurately convey a study’s true risks and benefits.^3^

Successful enrollments in acute stroke clinical trials require that research teams, including clinician investigators and study coordinators, are quickly notified and purposefully integrated into the clinical workflow. However, due to the emergency nature of cerebrovascular disease, clinician investigators, research coordinators, patients, and LARs, are often not in the same physical location when a potential trial candidate is identified, especially after hours.^1,4–6^ To overcome these physical barriers to enrollment, some programs utilize remote research practices (RRPs) in which ECTE processes are supported off-site.^1^ Hybrid remote and in-person clinical research has been proposed as a new standard for randomized control trials in order to address barriers related to cost, diversity, recruitment, and efficiency.^3^ Increasing enthusiasm for remote research has been tempered by the lack of evidence supporting RRP integration into real-world acute stroke trials. Consequently, skepticism regarding RRPs endures and adoption remains limited.

Our study formally compares the feasibility and effectiveness of RRPs to conventional in-person clinical research workflows for acute stroke clinical trials at two academic stroke programs.

## Methods

In this case-control study, we reviewed all screenings and enrollments at two academic stroke programs for two randomized, Phase-III, acute stroke intervention trials (MOST, NCT03735979 and TIMELESS, NCT03785678; June 19, 2019 to December 31, 2020) and an acute stroke observational study (MaRISS, NCT02072681; June 23, 2016-January 2, 2019) that required screening, informed consent, and enrollment within 24 hours. Both programs routinely incorporate RRPs utilizing telemedicine evaluation and telephone communication into clinical research operations across their clinical stroke networks. Remote research practices were defined as attempting completion of an ECTE without a member of the clinical research team present in-person. Study specific ECTEs for the following study elements: eligibility screening, informed consent, randomization, study intervention, inpatient study follow-up assessments, and outpatient study follow-up assessments are listed in Supplementary Table 1.

For each subject, prospectively maintained research databases (Medidata RAVE and WebDCU), the American Heart Association-Get With the Guidelines, and the electronic medical record were utilized to abstract demographic information. Individual subjects were further reviewed to determine: 1) the mode of clinical research (in-person, telemedicine evaluation, or telephone communication) used to complete each ECTE, 2) whether each ECTE was completed accurately and successfully, and 3) if a protocol violation occurred. When multiple research modalities were utilized the more “intensive” modality was assigned. For example, an informed consent discussion that was initiated by telephone or telemedicine and then continued in-person would be considered “in-person,” whereas an ECTE completed utilizing telephone communication and telemedicine would be considered ‘telemedicine’. Informed consent was successfully completed if the patient/LAR elected to participate in the study and the paper or electronic informed consent form (eConsent) was finalized and signed. Potential trial subjects who were clinically eligible, but who were not enrolled because they declined to participate or did not meet subsequent radiographic eligibility criteria were included in our assessment of eligibility screening and informed consent only. Likewise, not all follow-up study assessments were attempted on each enrolled subject. Early discharge, withdrawal from the study, or death obviated completion of subsequent inpatient or outpatient assessments, therefore, these were not required visits in the analyses.

At each site, the default clinical research workflow was to complete each ECTE in-person whenever possible, with the exception of randomization (routinely completed by an off-site coordinator focused exclusively on screening the electronic medical record and randomization). In complex study enrollments, RRPs were employed to overcome traditional barriers to enrollment. Telemedicine evaluation was used for clinical eligibility screening if subjects presented to spoke hospitals or arrived after hours when the clinical investigator and research coordinators were not present in the hospital. If the subject lacked the capacity to consent, eConsent forms were sent to a remotely located LAR and the informed consent discussion occurred via telephone or telemedicine video conferencing. Supervision of the study intervention could occur by phone or telemedicine when the clinical team was off-site. Inpatient and outpatient assessments were also preferentially completed in-person; however, outpatient follow-up via telephone and telemedicine was a common pre-existing practice for patients who did not reside locally.

The primary outcome was the rate of successful, accurate completion of ECTEs by research modality (in-person, telemedicine evaluation, telephone communication). The secondary outcome was the rate of protocol violations incurred by research modality. Fisher’s exact test was used to assess statistical significance for the primary and secondary outcome. If there was a significant difference between the three research modalities, head-to-head comparisons of each research modality were conducted using Fisher’s exact test. The rate of RRP utilization and indications for remote research were reported using descriptive statistics. The level of statistical significance was set at alpha = 0.05 and no corrections were made for multiple testing. All statistical analyses were completed using Stata (17.1, StataCorp LLC, College Station, TX).

### Clinical Studies

The TIMELESS trial is a prospective, randomized, double-blind clinical trial of acute ischemic stroke patients presenting with anterior circulation large vessel occlusion between 4.5-24 hours from last known well and NIHSS ≥ 5. To determine eligibility for randomization all participants underwent multimodal CT or MRI at baseline. After obtaining informed consent, participants with a target vessel occlusion and requisite core-penumbra mismatch were randomized to treatment with tenecteplase or placebo prior to endovascular thrombectomy. Administration of the study drug had to be completed prior to initiating endovascular thrombectomy or within 90 minutes of qualifying neuroimaging, whichever came first. Subjects then underwent inpatient and outpatient follow-up assessments culminating with the primary outcome: modified Rankin Scale (mRS) at 90 days.

The MOST trial is a prospective, single-blinded, multi-arm, randomized controlled trial studying acute ischemic stroke patients presenting with an NIHSS ≥ 6 who were treated with thrombolysis within three hours of their last known well time. Following eligibility screening and obtaining informed consent, subjects were randomized to adjuvant eptifibatide, argatroban, or placebo infusions. The study drug had to be initiated within one hour of thrombolysis as a three-minute bolus followed by a 0-2 hour infusion and 2-12 hour infusion with rate titrations dependent upon treatment arm and subsequent lab values. The infusion was complete after 12 hours. Subjects then underwent inpatient and outpatient follow-up assessments culminating with the primary outcome: mRS at 90 days.

The MaRISS study was an observational clinical study of patients presenting with acute ischemic stroke (confirmed on neuroimaging) within 4.5 hours and NIHSS ≤ 5 who did or did not receive intravenous alteplase. Patients underwent eligibility screening followed by informed consent within 24 hours. Subjects underwent inpatient and outpatient follow-up assessments culminating with the primary outcome: mRS at 90 days.

Institutional Review Board approval was obtained from each participating center to perform the study through a waiver of informed consent. De-identified data are available upon reasonable request to the corresponding author. The data is not publicly available to ensure privacy of research participants

## Results

One hundred and seventy-one consecutive patients who were screened for MaRISS, MOST, or TIMELESS were identified. Thirteen screened and consented patients did not meet the radiographic inclusion criteria and were ineligible for enrollment. Two additional patients were not enrolled: one patient’s NIHSS improved spontaneously, and one patient declined to participate. The remaining 156 eligible subjects were enrolled into three acute stroke trials: 99 were enrolled into an observational study of acute ischemic stroke and 57 patients were randomized into acute stroke intervention trials. Subject demographics were heterogenous and reflected the eligibility criteria of each clinical study (Table 1). All subjects were followed from the initial screening assessment until they completed the 90-day study, withdrew from the study, were lost to follow-up, or died.

**Table 1.** Baseline Demographics:

|  | **TIMELESS**<br>**(n=57)** | **MOST**<br>**(n=15)** | **MARISS**<br>**(n=99)** |
| --- | --- | --- | --- |
| NIHSS | 11 (8-22) | 13 (7-18) | 2 (1-4) |
| Age, median (IQR), years | 74 (64-83) | 73 (66-87) | 67 (58-74) |
| Sex (female) (n,%) | 30 (52.6%) | 9 (60.0%) | 45 (45.5%) |
| Race |  |  |  |
| -White (n,%) | 43 (75.4%) | 13 (86.7%) | 74 (74.7%) |
| -Asian (n,%) | 5 (8.8%) | 2 (13.3%) | 7 (7.1%) |
| -Black (n,%) | 5 (8.8%) | 0 (0%) | 16 (16.2%) |
| -Other or not reported (n,%) | 4 (7.0%) | 0 (0%) | 2 (2.0%) |
| Ethnicity |  |  |  |
| -Hispanic or Latino | 9 (15.8%) | 1 (6.7%) | 47 (47.5%) |
| Capacity to consent (yes) (n,%) | 6 (10.5%) | 3 (20.0%) | 77 (77.8%) |
| English speaker (yes) (n,%) | 50 (87.7%) | 14 (93.3%) | 65 (65.7%) |
| Patient/LAR required an interpreter for consent (n,%)* | 5 (8.8%) | 0 (0%) | 34 (34.3%) |

Abbreviations: IQR, interquartile range; LAR, legally authorized representative; n, number; NIHSS, NIH stroke scale
\*Does not match the number of English speaking patients because consent may have been obtained from an English-speaking LAR

### Remote Research Practice Utilization

A total of 1603 individual ECTE components were attempted: 739 (46.1%) in-person and 864 (53.9%) utilizing RRPs (309 [19.3%] telemedicine evaluation, 555 [34.6%] telephone communication) (Table 2). There was considerable heterogeneity between rates of RRPs versus in-person research across individual ECTEs (Figure 1). Randomization (94.7%) and outpatient study assessments (84.6%) were most commonly executed remotely, whereas eligibility screening (40.7%), informed consent (40.4%), supervision of the study intervention (44.6%), and inpatient assessments (18.8%) were less frequent. The primary reason(s) that remote research was utilized also varied between ECTEs. For time-critical ECTEs that occur early in an acute study enrollment, such as eligibility screening and informed consent, RRPs were most commonly utilized because the clinical investigator was remote from the subject. On the other hand, randomization (which was nearly universally completed remotely) and outpatient follow-up assessments primarily employed RRPs because the research coordinator was remote from the subject (Table 2).

**Table 2:**
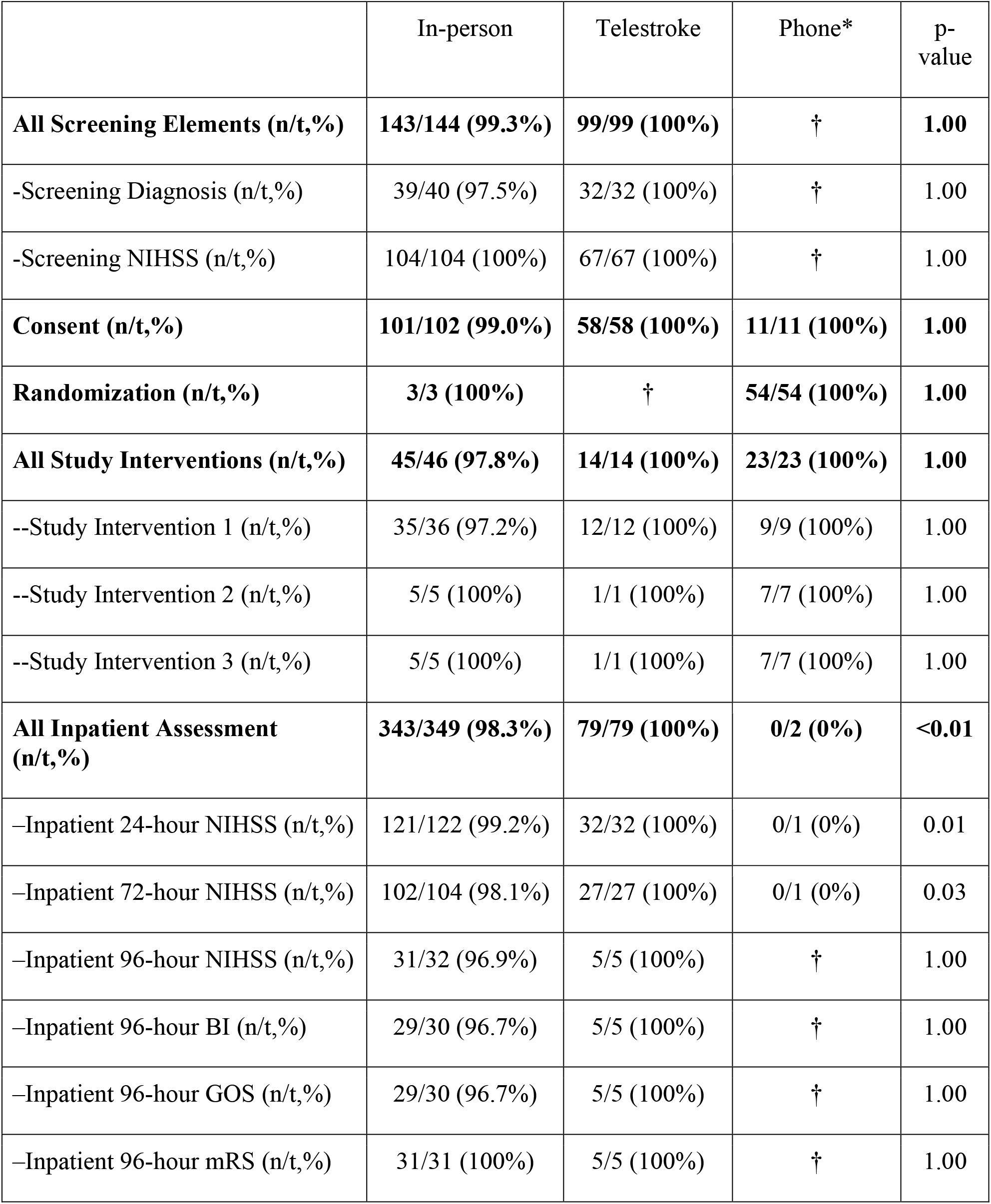
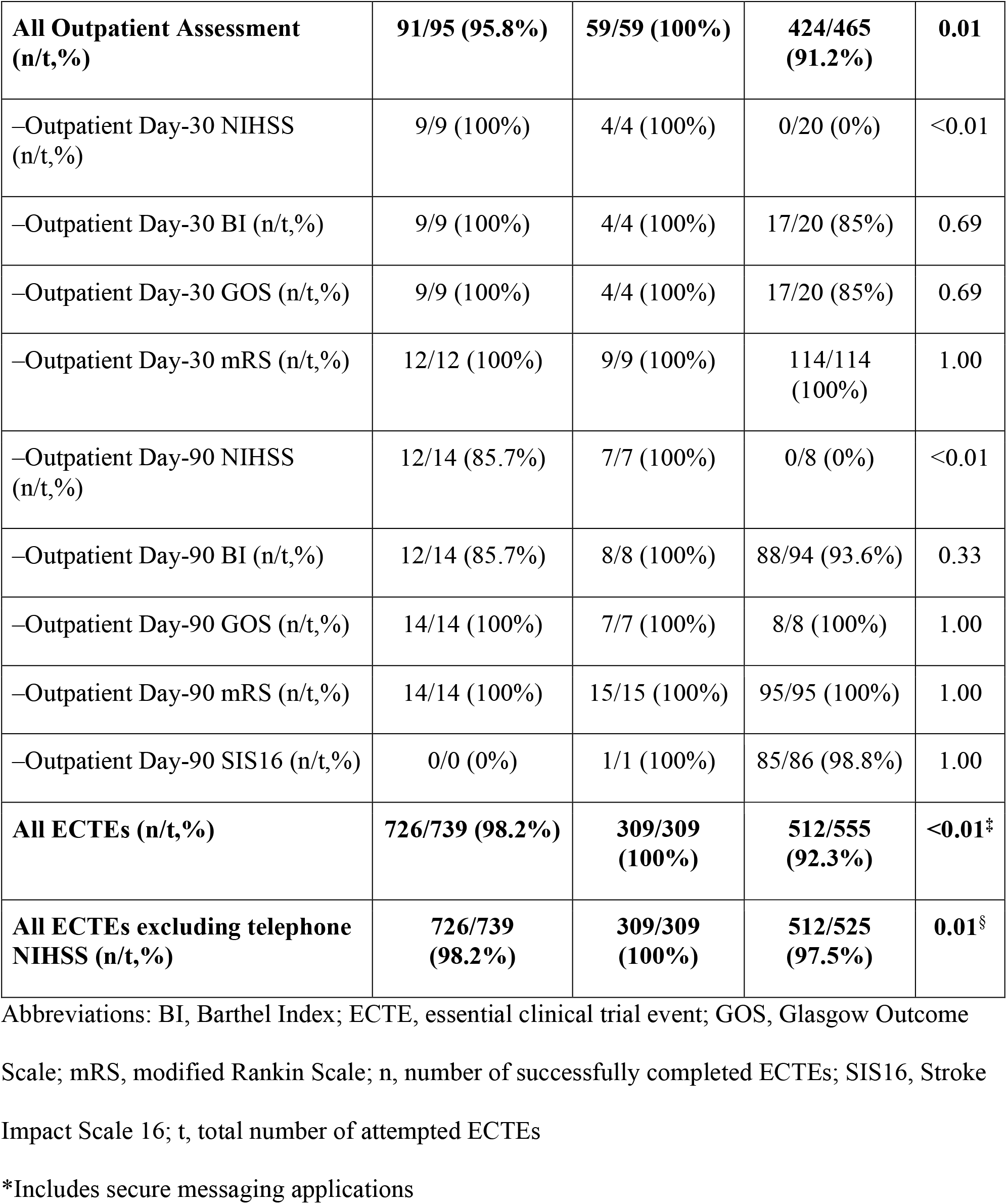
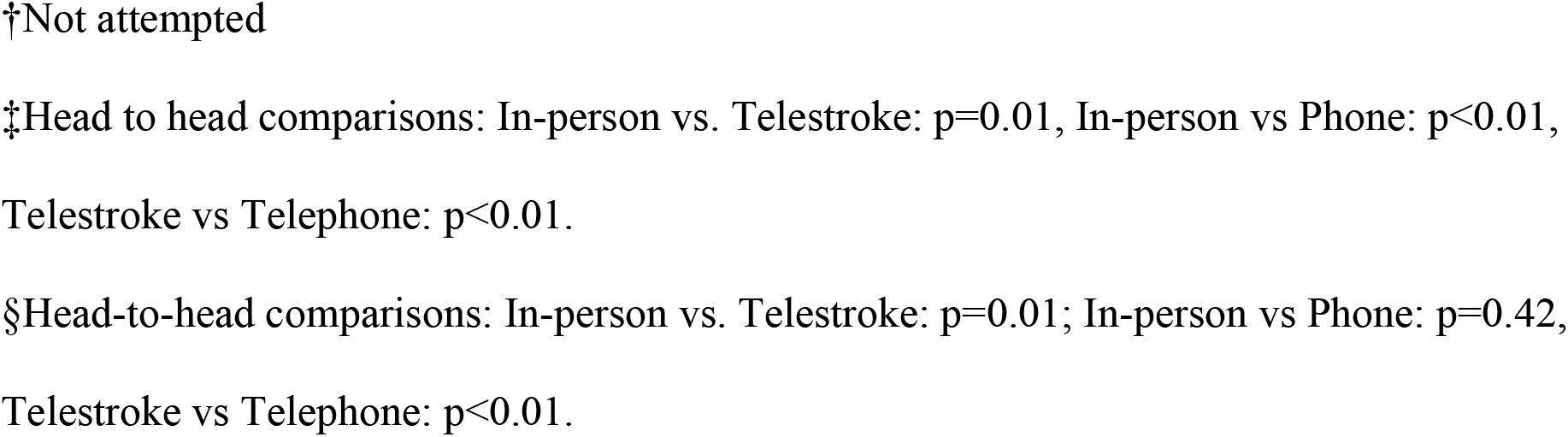
Successful Completion of ECTEs by Research Modality.

**Figure 1:**
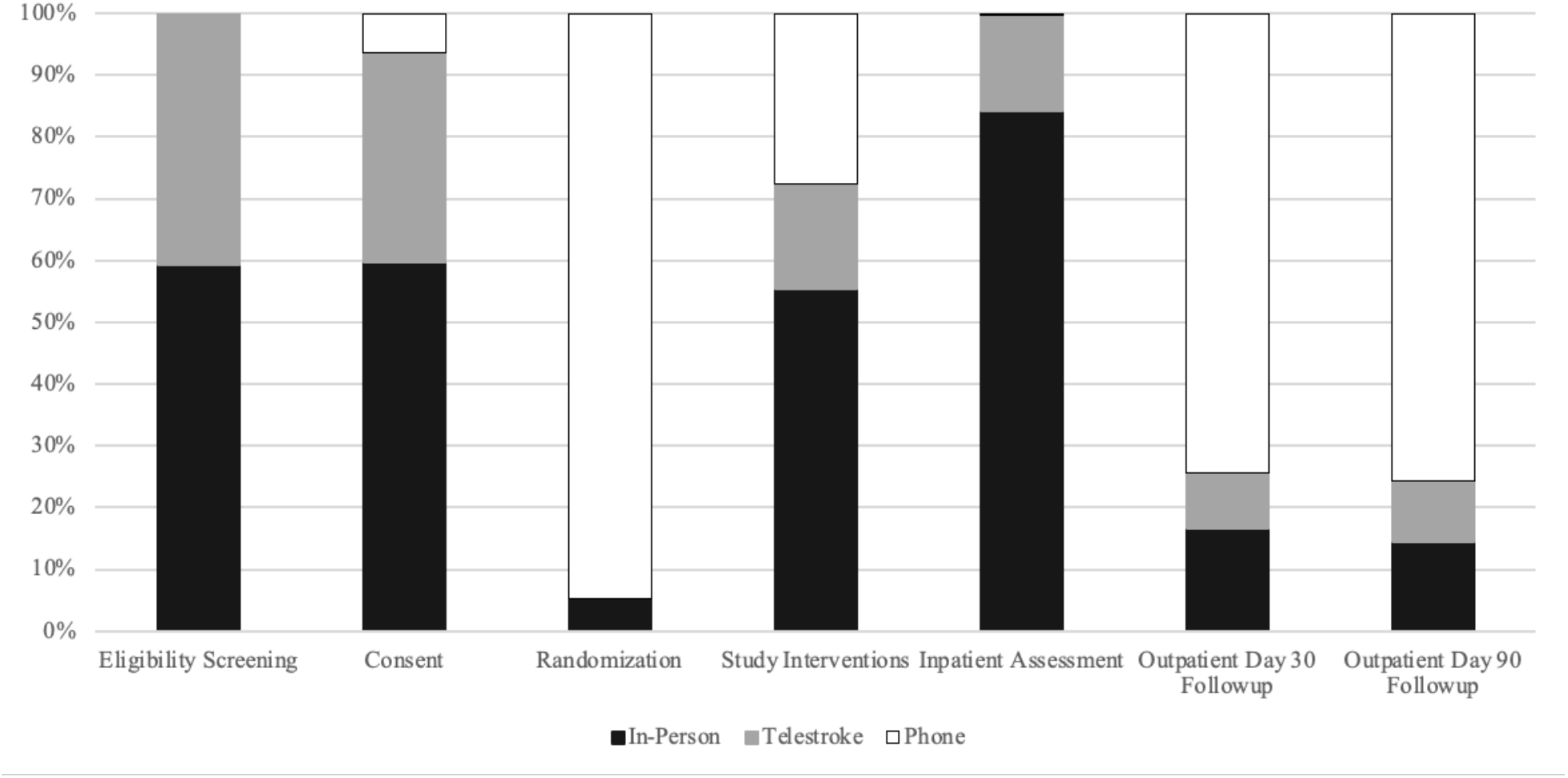
Research Modality Utilization By Essential Clinical Trial Event.

### Primary Outcome: Rate of Successful Completion of Essential Clinical Trial Elements by Research Modality

Of the 1603 individual ECTEs, we observed a significant difference in the rate of successful, accurate completion across research modalities: 98.2% in-person vs 100% telemedicine evaluation vs 92.3% by telephone communication (p <0.01). In subsequent head-to-head comparisons, ECTEs were significantly more likely to be completed successfully when executed via telemedicine evaluation compared to in-person research (p=0.01) and telephone communication (p<0.01). Additionally, ECTEs were more likely to be completed successfully when executed in-person than with telephone communication (p<0.01). These results were heavily impacted by the inability to obtain an NIHSS score via telephone follow-up. Although research teams were expected to obtain the follow-up NIHSS score, in recognition of barriers to subject retention (such as the COVID-19 pandemic), telephone follow-up was permitted and missed NIHSS outcome assessments were not considered a protocol violation. To account for this guidance, an analysis was also conducted excluding NIHSS assessments from the RRP telephone communication cohort. The successful completion rate of ECTEs by telephone communication rose to 97.5% and in-person execution of ECTEs was no longer superior to telephone (p=0.42), however, telemedicine evaluation remained superior to telephone communication (p<0.01). (Table 3)

**Table 3:** Primary Reason(s) Remote Research Practices were Utilized in Acute Stroke Clinical Trial Enrollments^*^.

|  | Primary Reason(s) for Utilizing Remote Research |  |  |  |  | Total Completed Using Remote Research |
| --- | --- | --- | --- | --- | --- | --- |
|  | MD remote | Patient remote | Coordinator Remote | COVID19 | Other |  |
| Eligibility Screening (n/t,%) | 64/67 (95.5%) | 35/67 (52.2%) | 13/67 (19.4%) | 9/67 (13.4%) | 1/67 (1.5%) | 67/171 (39.2%) |
| Informed Consent (n/t,%) | 62/69 (89.9%) | 43/69 (62.3%) | 12/69 (17.4%) | 8/69 (11.6%) | 1/69 (1.4%) | 69/171 (40.4%) |
| Randomization (n/t,%) | 8/54 (14.8%) | 2/54 (3.7%) | 35/54 (64.8%) | 10/54 (18.5%) | 0/54 (0%) | 54/57 (94.7%) |
| Supervision of Study Intervention (n/t,%) | 15/21 (71.4%) | 1/21 (4.8%) | 11/21 (52.4%) | 10/21 (47.6%) | 1/21 (4.8%) | 21/57 (36.8%) |
| Inpatient Study Assessments (n/t,%) | 32/34 (94.1%) | 28/34 (82.4%) | 2/34 (5.9%) | 5/34 (14.7%) | 0/34 (0%) | 34/155 (21.9%) |
| Outpatient Study Follow-up (n/t,%) | 22/234 (9.4%) | 87/234 (37.2%) | 201/234 (85.9%) | 28/234 (12.0%) | 4/234 (1.7%) | 234/259 (90.3%) |
| All Study Elements (n/t,%) | 203/479 (42.4%) | 196/479 (40.9%) | 274/479 (57.2%) | 70/479 (14.6%) | 7/479 (1.5%) | 479/870 (55.1%) |

Abbreviations: n, number of protocol deviations; t, total number of trial subjects completing the study element

### Secondary Outcome: Rate of Protocol Violations by Research Modality

A total of 870 consecutive study elements were assessed for protocol deviations; 391 (44.9%) were completed in-person, 194 (22.3%) over telemedicine, and 285 (32.8%) by telephone communication. We observed a significant difference in the rate of protocol deviations across research modalities (in person: 2.6%, telestroke 0.0%, and telephone: 2.8% [p=0.04]) (Table 4). In subsequent head-to-head comparisons, the rate of protocol deviations was significantly less with telestroke vs in-person (p=0.04) and telestroke vs telephone (p=0.02). There was no difference in the rate of protocol deviations between study elements completed in-person or via telephone (p=1.00). Subgroup analyses did not reveal significant differences in the rate of protocol deviations between individual study elements.

**Table 4:**
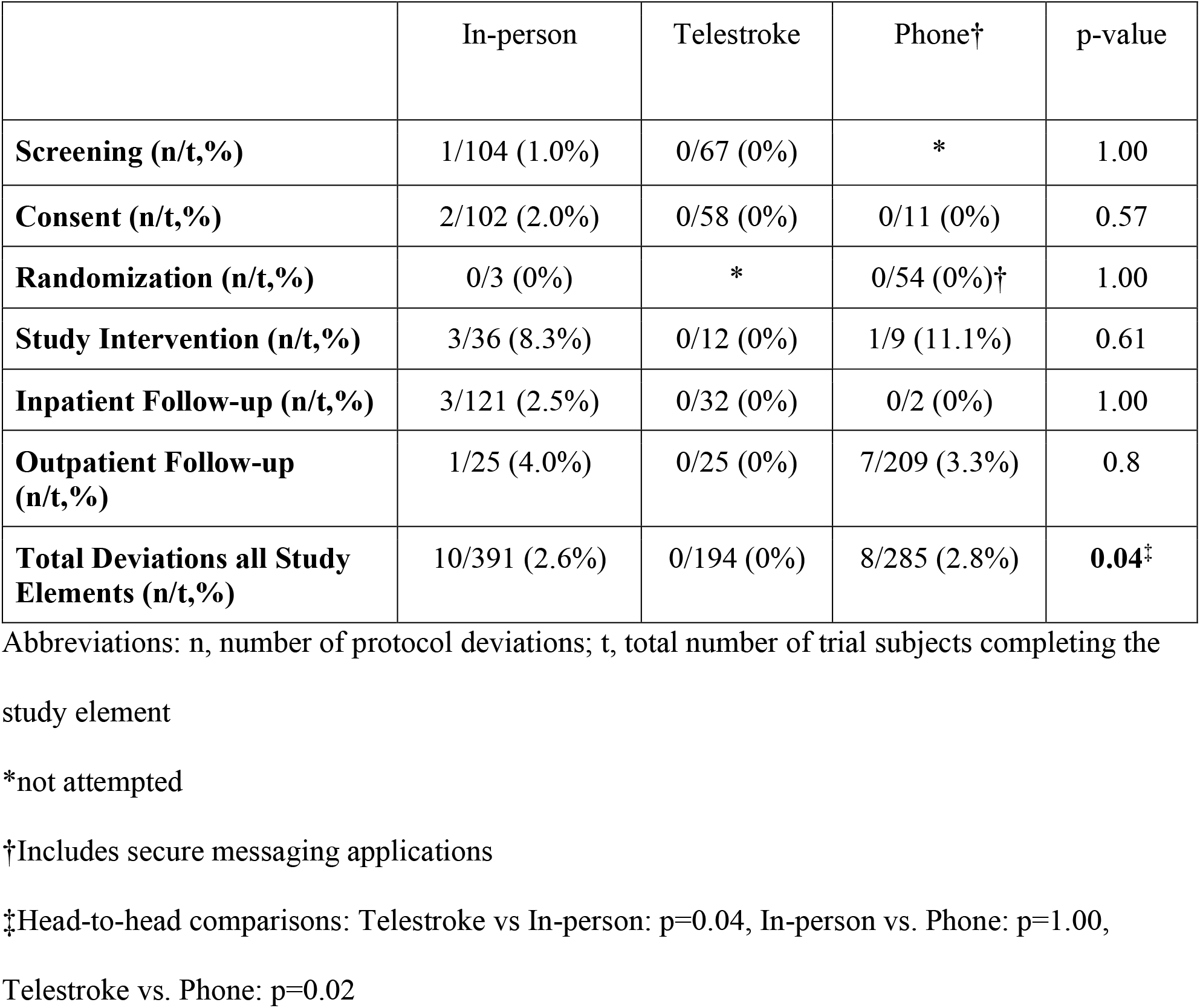
Protocol Deviations by Research Modality

|  | In-person | Telestroke | Phone† | p-value |
| --- | --- | --- | --- | --- |
| **Screening (n/t,%)** | 1/104 (1.0%) | 0/67 (0%) | * | 1.00 |
| **Consent (n/t,%)** | 2/102 (2.0%) | 0/58 (0%) | 0/11 (0%) | 0.57 |
| **Randomization (n/t,%)** | 0/3 (0%) | * | 0/54 (0%)† | 1.00 |
| **Study Intervention (n/t,%)** | 3/36 (8.3%) | 0/12 (0%) | 1/9 (11.1%) | 0.61 |
| **Inpatient Follow-up (n/t,%)** | 3/121 (2.5%) | 0/32 (0%) | 0/2 (0%) | 1.00 |
| **Outpatient Follow-up (n/t,%)** | 1/25 (4.0%) | 0/25 (0%) | 7/209 (3.3%) | 0.8 |
| **Total Deviations all Study Elements (n/t,%)** | 10/391 (2.6%) | 0/194 (0%) | 8/285 (2.8%) | **0.04‡** |

Abbreviations: n, number of protocol deviations; t, total number of trial subjects completing the study element
\*not attempted
†Includes secure messaging applications
‡Head-to-head comparisons: Telestroke vs In-person: p=0.04, In-person vs. Phone: p=1.00,
Telestroke vs. Phone: p=0.02

## Discussion

Although RRPs have the potential to address pervasive barriers to conducting high-quality clinical trials, including slow recruitment, limited patient access, lack of diversity, poor retention rates, and increasing economic costs, such promises remain largely theoretical.^1^ The real-world application of RRPs, especially in high-risk interventional stroke trials, remains unclear. In this study, ECTEs executed via telemedicine were actually completed more successfully when compared to conventional, in-person clinical research. The effectiveness of telemedicine in the acute stroke clinical trial setting should not be a surprise. Telemedicine already serves an analogous role in clinical practice: rapidly screening patients to assess eligibility for acute stroke treatment, obtaining informed consent for standard of care procedures, and conducting patient follow-up via telestroke are routine, widespread practices.^8,9^

Additionally, protocol deviations were less frequent when ECTEs were completed using telemedicine compared to in-person. This finding is especially salient: protocol deviations are a marker of adherence to study protocols that ensure the safety and autonomy of potential subjects, while delivering high-quality clinical data. Telemedicine RRPs could improve study protocol fidelity through streamlined, efficient clinical trial enrollments. Research teams may not be present in the emergency department when a potential subject arrives. By completing ECTEs remotely, time previously spent in transit can be repurposed towards trial execution thereby decompressing tight enrollment timelines. In fact, both sites in this study had already standardized off-site randomization due to perceived time savings. Even when not accounting for off-site randomization, more than half (53.4%) of all randomized subjects required RRPs to complete their ECTEs within the therapeutic window for trial enrollment (Figure 2). Thus, incorporation of RRPs doubled the number of possible enrollments in comparison to conventional research models that are restricted to in-person interaction alone.

**Figure 2:**
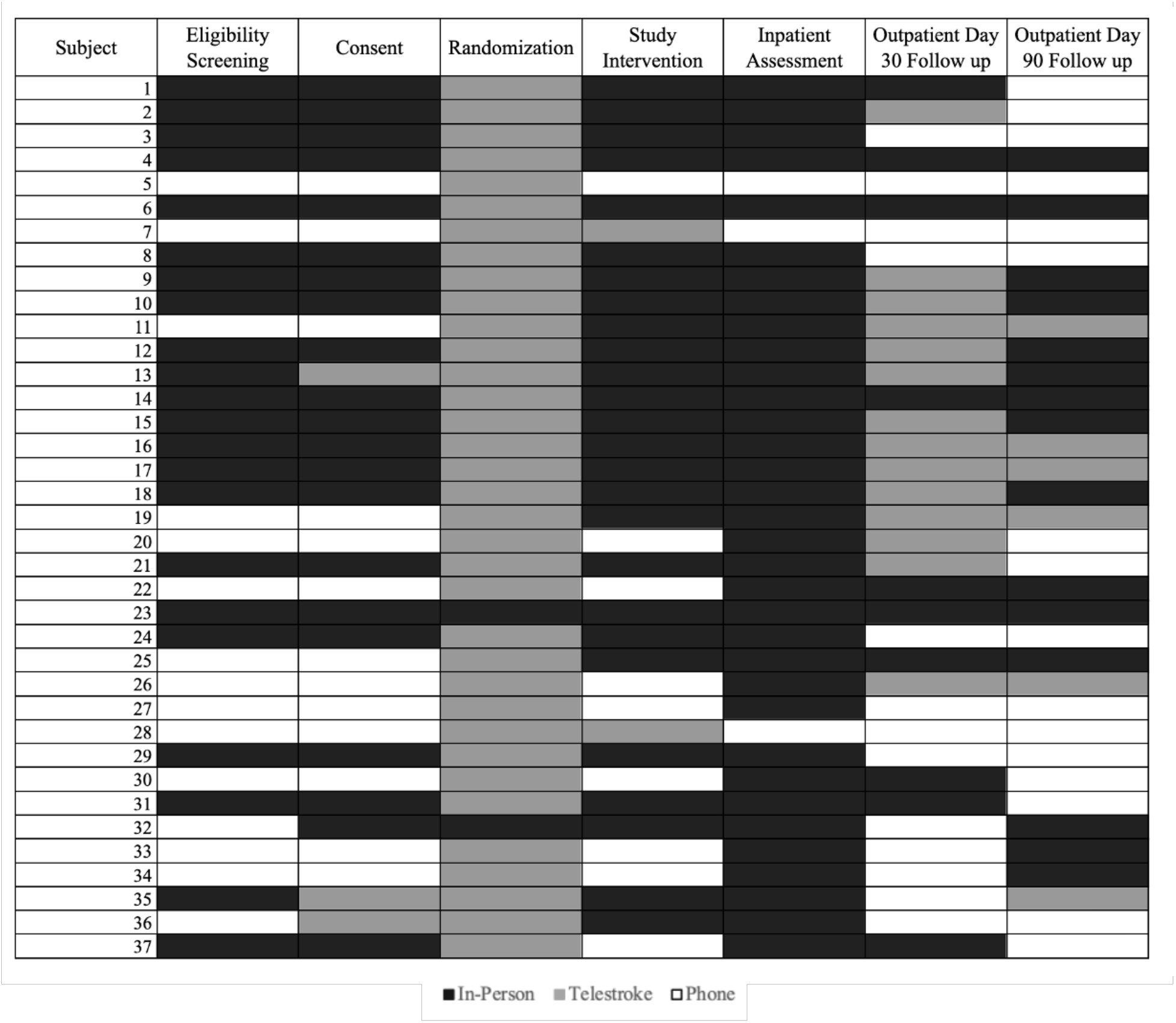
Research Modality Utilized for Each Study Element per Randomized Subject Who Completed the 90-day Enrollment.

In contrast to telemedicine evaluation, ECTEs executed via telephone communication were completed less successfully than in-person clinical research with a similar rate of protocol deviations. Moreover, telephone communication was not viable for initial clinical trial eligibility screening assessments although it has been validated for assessment of certain follow-up outcomes.^10^ These findings suggest that remote research conducted via telephone communication can have a role in acute stroke trial execution, but in a limited capacity.

Nonetheless, the overall high rates of successful ECTE completion and low rates of protocol deviations imply RRPs and in-person clinical research are complementary. Even within individual subjects, multiple research modalities (often all three) were typically employed during the 90-day study enrollment (Figure 2). The research modality utilized during the acute stroke trial enrollment was informed by clinical necessity. On the other hand, the modality of outpatient study follow-up was at the discretion of patients. Notably, the overwhelming majority of subjects elected to follow-up remotely via telestroke or telephone which is less physically and economically burdensome. Flexible execution of ECTEs with integrated RRPs allows for customizable clinical trial operations optimized for the clinical team, coordinators, and subject.^11^

This study demonstrated widespread, voluntary adoption of RRPs which were directly integrated into acute stroke clinical research workflows. Overall, 53.9% of all ECTEs were completed remotely. Three subjects completed their enrollment and follow-up solely utilizing RRPs without any in-person contact with the clinical research team (Figure 2, subjects 5, 7, 28). None of these patients incurred incomplete study assessments or protocol deviations during their 90-day enrollment. Additionally, 12 subjects (27.9%) enrolled in TIMELESS were randomized and received study intervention at affiliated stroke centers rather than the tertiary academic hospital. A further 28 (28.3%) enrollments in MaRISS occurred outside the tertiary academic stroke hospital. These enrollments hint at the broader potential of remotely supported research infrastructure to bring trials to eligible patients (rather than bringing patients to trials).

Decentralizing clinical research may facilitate more rapid and diverse recruitment while simultaneously advancing local clinical practice and ensuring that study findings are generalizable to the highest stroke-risk populations. ^3,4,10–12^ Remotely-supported research could also leverage economies of scale to expand the available hours of research coordinator support for acute trials.^1^ Unsurprisingly, 24-7 coverage is known to increase enrollment in comparison to coverage during working hours.^5,13^ Ultimately, RRPs open the possibility of streamlined, efficient clinical trial designs that maximize limited personnel and financial resources.^14^ Efficiency gains at the micro and macro level shorten clinical trial timelines accelerating meaningful clinical practice change.^3,6^

Our findings are retrospective and may be confounded by selection bias or indication. Telemedicine follow-up and eConsent require technical competency and access to the internet. Subjects utilizing RRPs may represent a cohort with increased economic resources and education who are more likely to complete all follow-up assessments. On the other hand, RRPs were only utilized when physical distance separated the clinical team, patient, LAR, or subject. These inherently more complex research settings could have negatively impacted the rate of ECTE completion and protocol deviations in the telemedicine and telephone cohorts. Additionally, we were unable to assess the impact of RRPs on patients’ willingness to participate in clinical trials. Both sites introduce clinical trials early in the evaluation of acute stroke patients before initiating formal informed consent. Patients who declined clinical trial participation upfront would not interface with the research coordinator team or be included in screening logs. Finally, these findings may not be generalizable across different hospital systems. Remote research workflows require ongoing training, coordination, and investment from all involved parties;^15^ on-site assistance with study drug delivery, administration of the study intervention, and patient monitoring is still essential. Both stroke programs in this study have considerable accumulated trial experience that extends to bedside nursing and pharmacy staff which may enable effective RRPs.

## Conclusion

Remote research practices were effective and doubled randomized ASCT enrollments in comparison to conventional research models that are restricted to in-person interaction alone. Telemedicine was associated with the highest rate of successful ECTE execution and the lowest rate of protocol deviations. These findings may be confounded by indication and further definitive study is indicated.

## Data Availability

N/A

N/A

## Acknowledgments

None

